# Identifying modifiable comorbidities of schizophrenia by integrating electronic health records and polygenic risk

**DOI:** 10.1101/2023.06.01.23290057

**Authors:** Tess Vessels, Nicholas Strayer, Karmel W. Choi, Hyunjoon Lee, Siwei Zhang, Lide Han, Theodore J. Morley, Jordan W. Smoller, Yaomin Xu, Douglas M. Ruderfer

**Author notes:** **Corresponding authors:** Douglas M. Ruderfer, Yaomin Xu.

## Abstract

Patients with schizophrenia have substantial comorbidity contributing to reduced life expectancy of 10-20 years. Identifying which comorbidities might be modifiable could improve rates of premature mortality in this population. We hypothesize that conditions that frequently co-occur but lack shared genetic risk with schizophrenia are more likely to be products of treatment, behavior, or environmental factors and therefore potentially modifiable. To test this hypothesis, we calculated phenome-wide comorbidity from electronic health records (EHR) in 250,000 patients in each of two independent health care institutions (Vanderbilt University Medical Center and Mass General Brigham) and association with schizophrenia polygenic risk scores (PRS) across the same phenotypes (phecodes) in linked biobanks. Comorbidity with schizophrenia was significantly correlated across institutions (r = 0.85) and consistent with prior literature. After multiple test correction, there were 77 significant phecodes comorbid with schizophrenia. Overall, comorbidity and PRS association were highly correlated (r = 0.55, p = 1.29×10^−118^), however, 36 of the EHR identified comorbidities had significantly equivalent schizophrenia PRS distributions between cases and controls. Fifteen of these lacked any PRS association and were enriched for phenotypes known to be side effects of antipsychotic medications (e.g., “movement disorders”, “convulsions”, “tachycardia”) or other schizophrenia related factors such as from smoking (“bronchitis”) or reduced hygiene (e.g., “diseases of the nail”) highlighting the validity of this approach. Other phenotypes implicated by this approach where the contribution from shared common genetic risk with schizophrenia was minimal included tobacco use disorder, diabetes, and dementia. This work demonstrates the consistency and robustness of EHR-based schizophrenia comorbidities across independent institutions and with the existing literature. It identifies comorbidities with an absence of shared genetic risk indicating other causes that might be more modifiable and where further study of causal pathways could improve outcomes for patients.

## Introduction

Patients with schizophrenia have a reduced life expectancy of 10-20 years.^1,2–4^ Contributing factors include smoking, substance abuse, medication side effects, poor health maintenance, accidents, and suicide.^2,3,5^ However, medical comorbidities account for 70% of the premature mortality experienced by schizophrenia patients.^6,7^ The mortality ratio for schizophrenia increases almost four-fold when comorbid with other conditions^4,8^ and continues to increase with additional comorbidities.^4^ Multimorbidity (two or more comorbidities) is common, occurring in 64% of schizophrenia patients.^6^ Underdiagnosis of comorbid conditions and delays in seeking medical care are important factors contributing to early mortality.^7^ Schizophrenia has been associated with a wide variety of comorbidities across most major organ systems such as cancer, endocrine, nutritional and metabolic diseases, respiratory diseases, cardiovascular diseases, digestive system diseases, neurological disorders, musculoskeletal diseases, genitourinary diseases, and immunological diseases.^2,6,9–32^

Schizophrenia is a highly heritable disorder that shares significant genetic risk with multiple other phenotypes.^33,34^ A recent study combined data from four independent health care systems to assess the relationship between genetic risk of schizophrenia and other phenotypes.^35^ Using a schizophrenia polygenic risk score (PRS), they performed a phenome-wide association study (PheWAS) at each site and combined the results in a cross-site meta-analysis. The study identified associations of the schizophrenia PRS with psychiatric conditions, neurological disorders, obesity, urinary syndromes, viral hepatitis, synovitis and tenosynovitis, and malaise and fatigue.^35^ Genetic risk of schizophrenia has also recently been linked with poor cardiac phenotypes.^36^ These studies demonstrate the power of biobanks linked to EHRs for identifying phenotypes having shared genetic risk with schizophrenia.

Comorbidity or phenotypic correlation is often a byproduct of shared underlying genetic risk or genetic correlation.^37^ Therefore, it is expected that most schizophrenia comorbidities will share genetic risk with schizophrenia. Of particular importance, are schizophrenia comorbidities that do not share any underlying genetic risk with schizophrenia. These comorbidities are more likely to be driven by adverse events from treatment, unhealthy behavior, or environmental factors as opposed to innate biological risk and therefore are more likely to be modifiable.^38^ Identifying this set of comorbidities could lead to improved causal understanding and preventative efforts to reduce these risks. Leveraging EHR data linked to biobanks can provide a direct path to identifying and comparing comorbidities with and without genetic association to implicate potentially modifiable comorbidities.

Here, we assessed comorbidity using EHR billing codes, adjusting for demographic and health care utilization covariates in two health care systems, and validated our findings with prior literature. Associations of schizophrenia PRS with the same set of codes across two linked biobanks were used to identify phenotypes with shared schizophrenia genetic risk. We then applied equivalence testing to identify comorbidities that had significant absence of schizophrenia PRS association. These phenotypes represent opportunities to identify other, potentially modifiable causes of these comorbidities.

## Methods

### Quantifying the degree of comorbidity with schizophrenia using phecodes from two health care systems

Demographic information including age, sex, ethnicity, and International Classification of Diseases (ICD) billing codes were obtained for 250,000 randomly chosen people from the EHRs of two major health care systems, Vanderbilt University Medical Center (VUMC) and Mass General Brigham (MGB). MGB included both ICD version 9 and 10 while VUMC used ICD-9 only. We selected 250,000 patients to balance a representative sample and computing burden. The createPhenotypes() function from the PheWAS package in R was used to consolidate the ICD codes and their counts into broader clinical descriptions known as phecodes (definition 1.2).^39–41^ Phecodes listed as exclusions (conditions that are similar to the phecode of interest) were removed from the controls (https://phewascatalog.org/phecodes).^42^ The method is explained in further detail here.^43^

Pairwise logistic regressions were performed on the set of patients for each health care system separately across all phecodes, of which 1,701 phecodes occurred in at least one patient with schizophrenia. A person was considered a case for a phenotype if two or more phecodes were present and a control if no phecodes for that phenotype were present. Patients with a single instance of a phecode were considered inconclusive and treated as missing in the regressions. Covariates included were current age, age at last visit (EHR age), sex (male, female, or unknown), inferred or self-reported race (Asian, Black, Native American/Alaskan, Other, Pacific Islander, Unknown, or White), and the log of the number of unique phecodes per patient to account for healthcare utilization. The results from the regressions at each health care system were then combined using the average of Z-scores weighted by the number of people with both phecodes at their corresponding institutions. The p-values for these clinical associations were estimated using these weighted Z-scores and the following formula, P = 2*pnorm(-abs(Z-score)). The P-values were then Bonferroni corrected for the number of phecodes using the p.adjust() function in R.

### Schizophrenia comorbidity literature review

We searched the published literature as of September 15, 2021, for previously identified comorbidities of schizophrenia. We used medical subject headings or MeSH terms “schizophrenia” and “comorbidity” or “multimorbidity” to search PubMed for relevant papers. The search was restricted to papers from the past ten years where the full text was available in English, resulting in 866 papers. We retained only case/control studies, retrospective cohort studies, review articles, and meta-analyses, removing 99 papers that were clinical trials, case reports, or letters to the editor. Requiring that the papers detailed comorbid relationships with individual disorders excluded an additional 727 papers. These papers often addressed comorbidities as a group among specific circumstances (e.g., COVID-19 pandemic, imaging data for volumetric differences in gray matter, post-surgery period, or healthcare utilization). In total, 40 papers remained, and the results section was reviewed for any mention of a comorbid relationship with schizophrenia. Among this set, the phenotypes mentioned in at least 2 papers were included. There was a total of 64 comorbidities for schizophrenia from these papers of which 53 mapped to a unique phecode. The other 11 comorbidities did not map for lack of or too much specificity (e.g., cancer vs. malignant neoplasm of thoracic esophagus). Since these phenotypes were gathered across multiple papers, they do not have a comparable quantitative metric to represent their effect sizes. A hypergeometric test was performed to determine if the overlap of literature-based comorbidities and EHR-based comorbidities was greater than expected by chance.

### Genotype sample description and quality control (QC)

Genotyping quality control (QC) and imputation were performed individually at both institutes. Similar filtering steps were applied to both genotype sets such as excluding single nucleotide polymorphisms (SNPs) based on missingness rate, minor allele frequency (MAF), Hardy-Weinberg Equilibrium (HWE), and strand ambiguity. Individuals were removed based on sex discordance, excess heterozygosity, or relatedness. Detailed sample descriptions and QC are provided for each site separately in the Supplementary Methods.

### PRS calculation and PheWAS

PRS were calculated using the Python based program PRS-CS.^44^ PRS-CS is a Bayesian regression approach that uses continuous shrinkage priors for SNP effect sizes and the 1000 Genomes Project European cohort to represent the linkage disequilibrium (LD) structure between SNPs.^44^ The SNP effect sizes from the Psychiatric Genomic Consortium’s (PGC) latest available genome-wide association study (GWAS) for schizophrenia were used to calculate the PRS.^45^ These results came from a meta-analysis of 90 cohorts that included 67,390 cases and 94,015 controls of European and East Asian ancestries as well as 7,386 cases and 7,008 controls for African American and Latino ancestries, resulting in 263 genome-wide significant (GWS) loci from the initial 7,585,078 SNPs analyzed.^45^ For European ancestries specifically, there were 53,386 cases and 77,258 controls.^45^ The scaling parameter, phi, was set to 1e-2 to represent the polygenic architecture of schizophrenia. The PRS-CS generated posterior effects for SNPs were then used to calculate PRS for each cohort using the PLINK1.9–score flag with sum modification to output SCORESUM instead of SCORE.^46^ PRS PheWAS analysis was conducted two ways: with schizophrenia patients included and with them excluded. PRS PheWAS with schizophrenia patients excluded was ultimately used to reduce bias from schizophrenia cases.

For each of the phecodes with at least one person as a case (N =1,768 VUMC; N=1,639 MGB; N=1,586 Combined), we performed a logistic regression testing its association with the schizophrenia PRS including covariates of sex, current age, and the first 10 principal components (PCs) of ancestry within the European population where sample size was sufficient. As proxy variables for healthcare utilization and morbidity, we included record length in days as a covariate. As above, at least two instances of the phecode had to be present to be considered a case and phecodes listed as exclusions were removed from the controls. The cross-site information was combined in a fixed-effect inverse variance-weighted meta-analysis using the phewasMeta() function included in the PheWAS package.^40^

### Identification of comorbid phenotypes with significant lack of PRS effect

An equivalence test was used to assign statistical significance to how similar the distributions of schizophrenia PRS were between cases and controls for each of the 77 significant EHR comorbidities. The tost() function from the TOSTER package version 0.4.1 in R was used to perform two one-sided t-tests (TOST) of the PRS between cases and controls.^47^ A Cohen’s d of 0.2 and -0.2 were used as the equivalence bounds. The TOST test was performed at each site independently. The P-values from the TOST test were Bonferroni corrected for multiple comparisons based on the number of phecodes tested at both sites and with meta-analysis PRS results (N=65). These phenotypes were meta-analyzed according to Fisher’s method for combining p-values to estimate chi squared values and then converted to p-values using the pchisq() function in R and then applying Bonferroni correction for the shared 65 phenotypes. To isolate phecodes that did not share genetic risk with schizophrenia, we reduced this set of phenotypes to include only those that were non-significant for the meta-analysis of the schizophrenia PRS.

## Results

### Defining EHR-based comorbidities of schizophrenia

Pairwise logistic regressions of billing codes grouped into 1,701 phecodes were performed accounting for demographic and utilization information on clinical data from two health care systems (VUMC and MGB) to quantify comorbidity (see Methods).^43^ For all pairs of phecodes, we observed significant correlation (r = 0.79, Pearson) of comorbidity across sites. For comorbidity with schizophrenia, the correlation across sites was stronger (r = 0.85, Pearson). Given the consistency across institutions, we defined our primary comorbidity measure as the weighted average of the Z-scores across VUMC and MGB.

After Bonferroni correction for 1,701 tests (p < 2.94×10^−5^), there were 77 significant phenotypes that were comorbid with schizophrenia (Figure 1). Forty-six of these were psychiatric phenotypes including “Psychosis”, “Suicidal ideation”, “Bipolar disorder”, “Suicidal ideation or attempt”, “Conduct disorders”, “Mood disorders”, and “Hallucinations”. Thirty-one were other medical codes including seven neurological: “Epilepsy, recurrent seizures, convulsions”, “Torsion dystonia”, “Convulsions”, “Extrapyramidal disease and abnormal movement disorders”, “Chronic pain”, “Epilepsy”, and “Other and unspecified disorders of the nervous system”. The second largest group of non-psychiatric comorbidities was injuries and poisonings: “Poisoning by psychotropic agents”, “Poisoning by anticonvulsants and anti-Parkinsonism drugs”, “Adverse drug events and drug allergies”, “Toxic effect of other substances, chiefly nonmedicinal as to source”, “Poisoning by drugs primarily affecting the autonomic nervous system”, and “Skull and face fracture and other intercranial injury”. A complete list of the significant phenotypes for the combined, VUMC only, and MGB only EHR comorbidities are reported in Supplementary Table 1 and Supplementary Figure 1 and 2.

**Figure 1:**
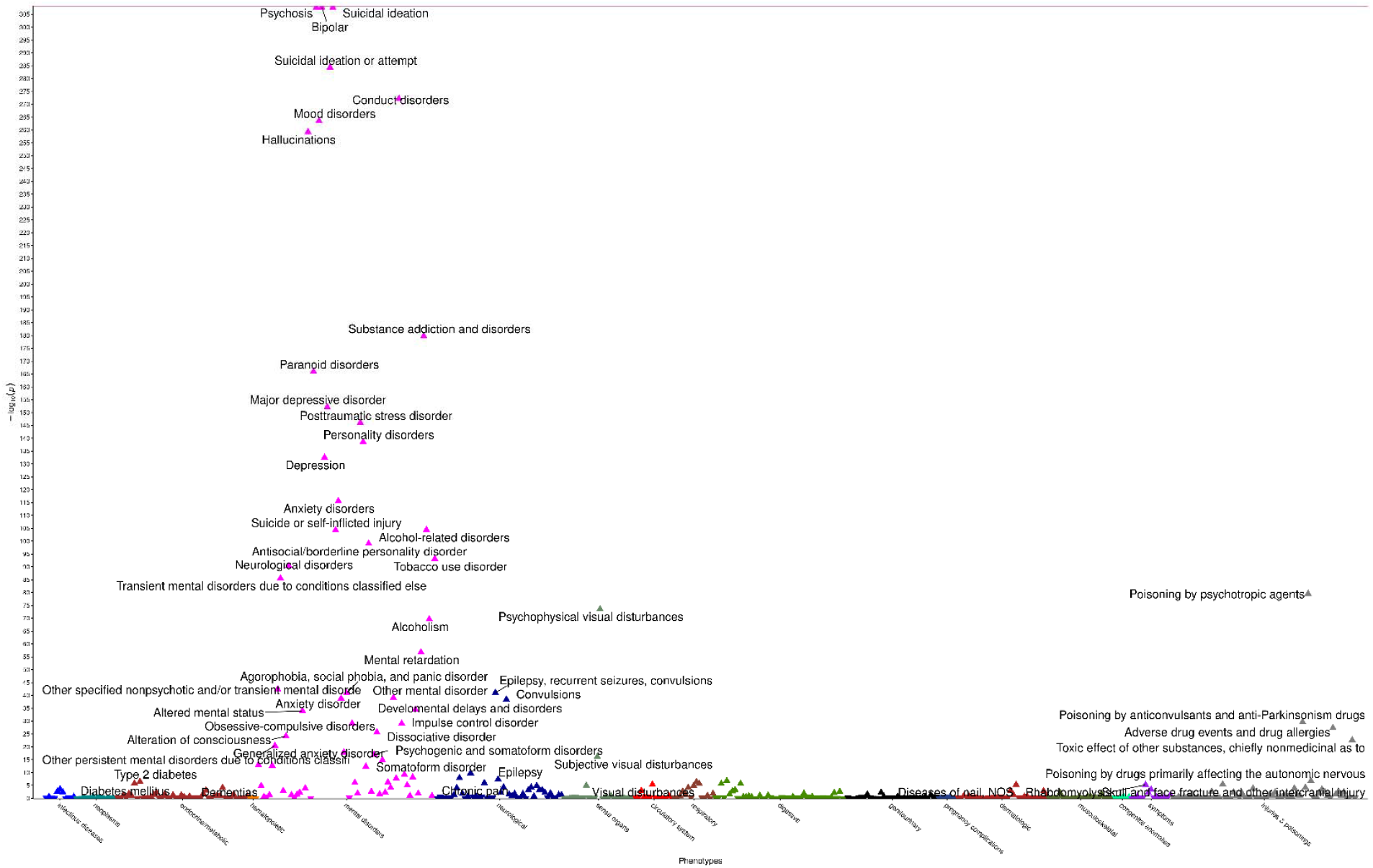
Bonferroni significant EHR-based schizophrenia comorbidities across both institutes. The x-axis is phecodes grouped by category. The y-axis is the negative log10 of the p-values for the phenotypes associated with schizophrenia from the regressions. Labeled triangles represent phenotypes that showed significant association after Bonferroni correction, with either positive (upright triangle) or negative (inverted triangle) direction of effect.

### EHR based comorbidities show consistency with comorbidities described in the literature

To assess the robustness of the EHR-based schizophrenia comorbidities, we compared these results to those described in the literature. A comprehensive literature review identified 40 papers implicating specific phenotypes significantly comorbid with schizophrenia. Among these papers, there were 38 medical and 17 psychiatric comorbidities that were identified in 2 or more of the papers and matched to the closest specific phecodes in our data (Table 1). Among the 38 medical phenotypes, 7 were significant and had a positive effect size in our EHR data. Of the 17 psychiatric phenotypes, 16 were significant with a positive effect size in our data. When combined, 23 out of 55 comorbidities replicated based on EHR comorbidities which was significantly (p=2.77×10^−11^) more than expected due to chance based on a hypergeometric test. Overlap among the medical phenotypes (p = 4.11×10^−3^) and the psychiatric phenotypes (p = 6.07×10^−24^) were also independently significant. There were 47 phenotypes significantly associated with schizophrenia in the EHR but without multiple mentions across the papers. Most were among the psychiatric, neurological, and injuries and poisoning categories (Supplementary Table 2).

**Table 1:**
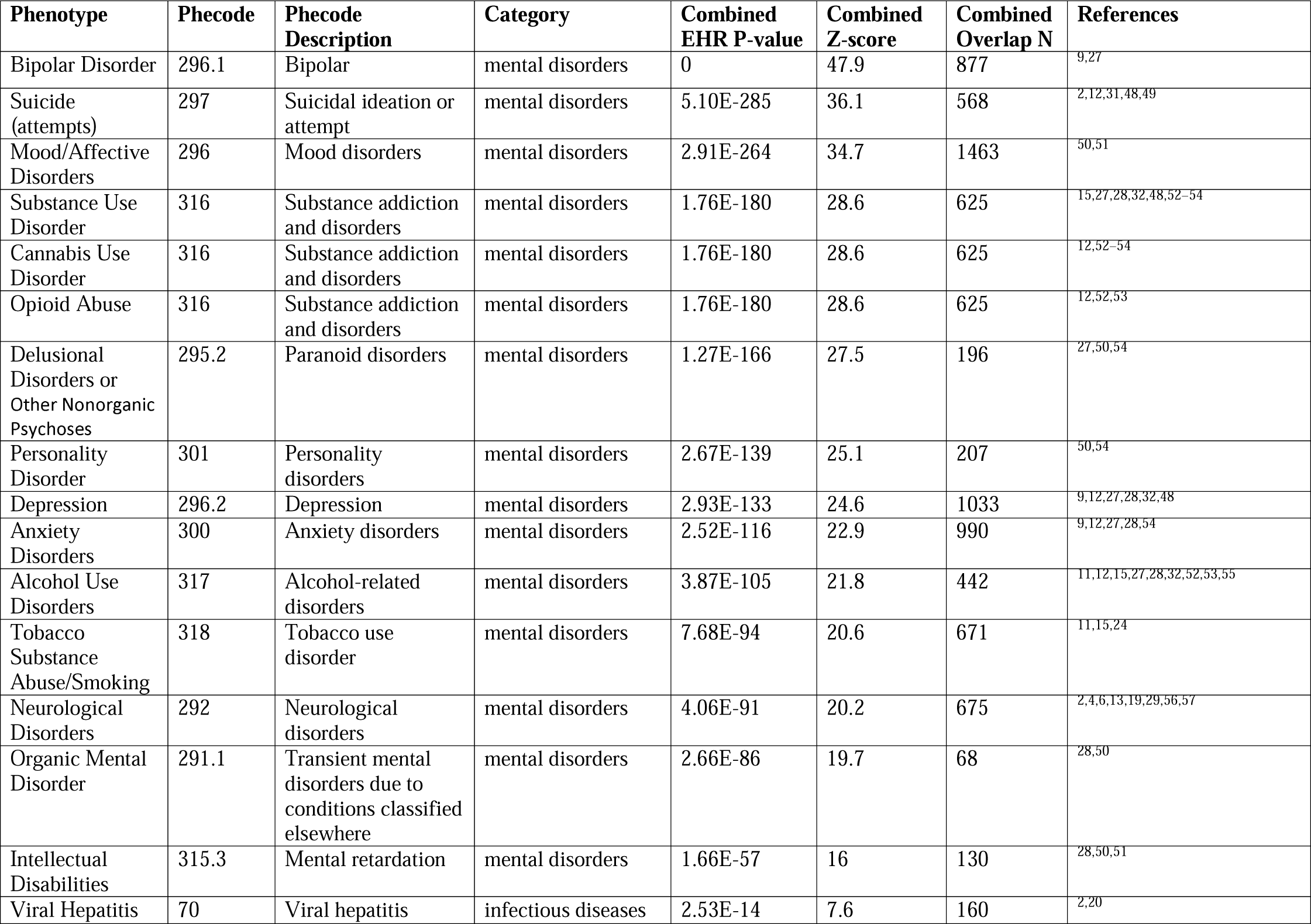
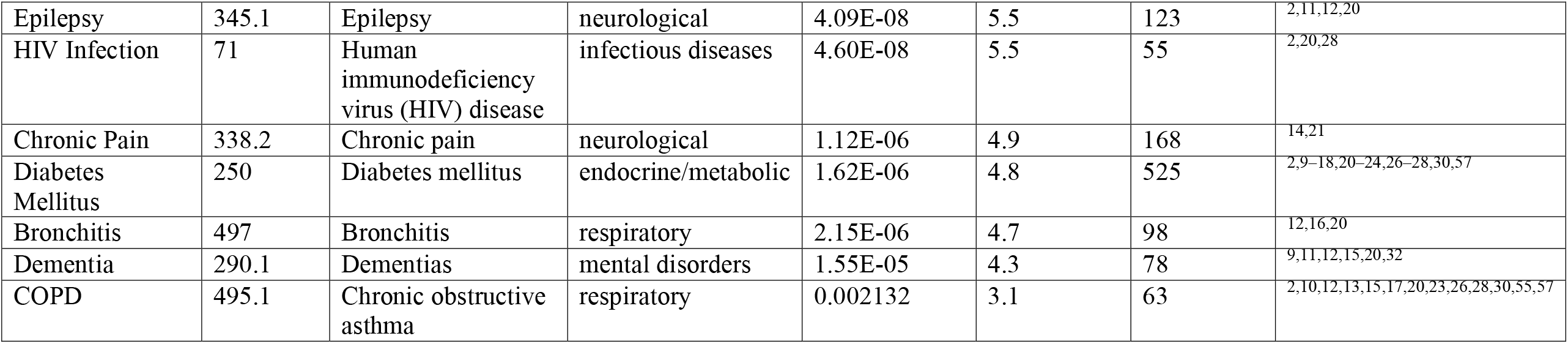
Comparison of schizophrenia comorbidities in the literature to our EHR-based comorbidities.

### Identifying phenotypes associated with genetic risk of schizophrenia

We next sought to identify which phecodes were significantly associated with schizophrenia PRS among patients of European ancestries (Supplementary Table 3, Supplementary Figures 3-4), where we had sufficient sample size, in biobanks at VUMC (N=64,190) and MGB (N=25,698). Schizophrenia cases were removed to exclude association as a byproduct of comorbidity. Results were combined using a fixed-effect inverse variance-weighted meta-analysis (see Methods). After Bonferroni correction for 1586 tests (p < 3.15×10^−5^), there were 38 significant schizophrenia PRS-associated phenotypes (Figure 2). As with the EHR comorbidities, the majority were psychiatric conditions such as “Bipolar disorder”, “Anxiety disorder”, and “Suicidal ideation”. There were also a variety of medical illness associations such as “Lice infestation”, “Dysuria”, “Viral hepatitis C”, “HIV”, and “Poisoning by psychotropic agents”. Schizophrenia PRS associations were also conducted for the AFR ancestry sample, but no significant associations were identified (Supplementary Figure 5-6).

**Figure 2:**
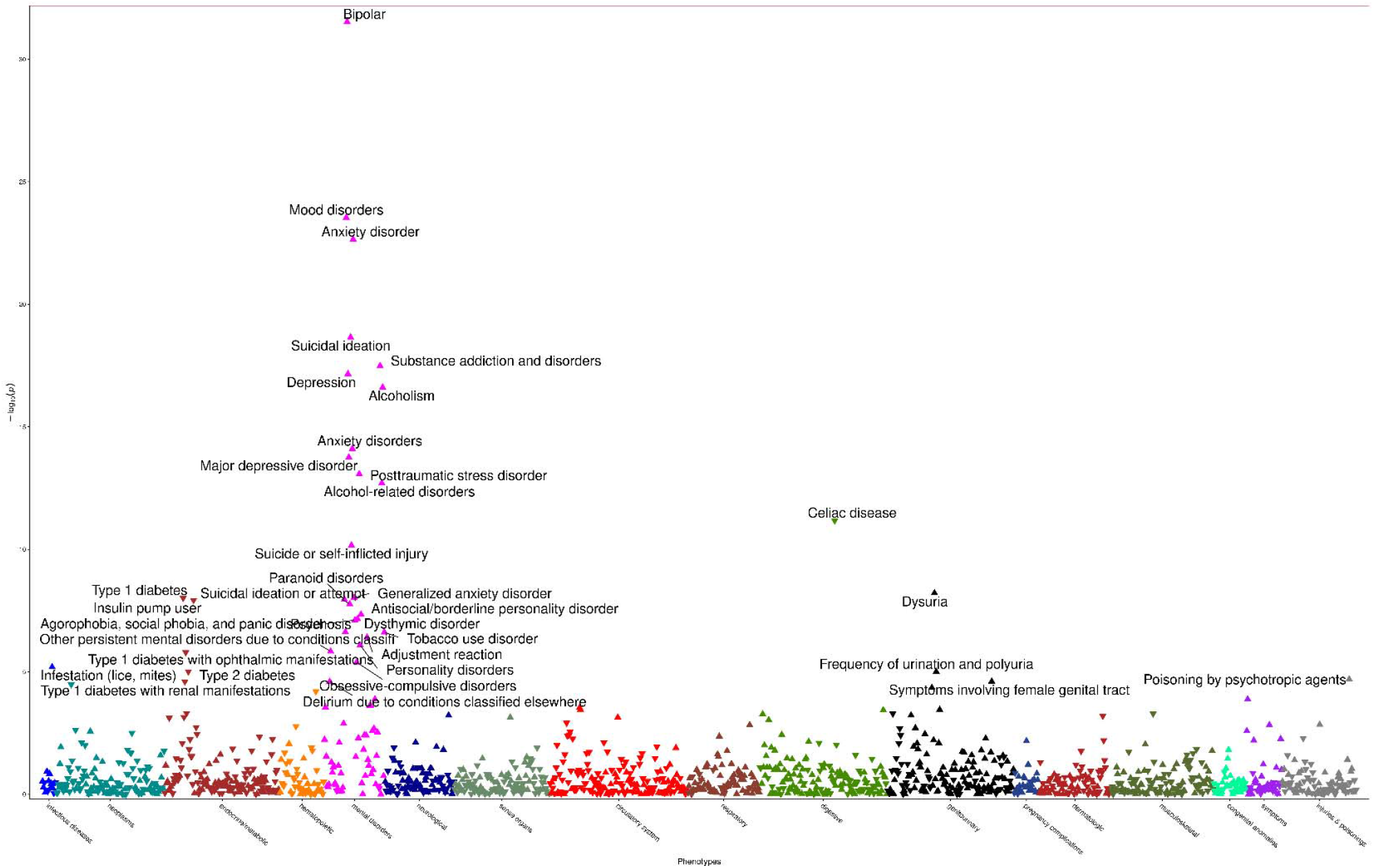
Bonferroni significant schizophrenia PRS meta-analysis associations. The x-axis is phecodes grouped by category. The y-axis is the negative log10 of the p-values for the phenotypes associated with schizophrenia PRS. Labeled triangles represent phenotypes that showed significant association after Bonferroni correction, with either positive (upright triangle) or negative (inverted triangle) direction of effect.

### Identification of comorbidities with significant absence of genetic contribution

Overall, the EHR-based comorbidities were significantly correlated with the PRS associations (r = 0.55, p = 1.29×10^−118^). Similar correlations were seen when comorbidities and PRS associations were compared across sites to remove sample dependence: VUMC comorbidities with the MGB PRS associations (r = 0.46, p= 8.56×10^−77^) and the MGB comorbidities with the VUMC PRS associations (r = 0.46, p = 5.31×10^−68^). We were primarily interested in identifying phecodes with significant comorbidity but a lack of shared genetic overlap with schizophrenia. These phenotypes represent those for which common genetic variation is not a strong driver of comorbidity, implicating other potential contributors such as environmental influences, medication effects or rare genetic variation.^38^ For each of the 65 significant EHR comorbidities that also had PRS results, the distributions of PRS for cases and controls were compared using a TOST equivalence test to determine whether they were significantly equivalent to each other (i.e., significant absence of difference in PRS between cases and controls, see Methods). After multiple test correction, 36 comorbidities were considered significantly equivalent (Table 2 and Supplementary Table 4). Included in this set were 11 phenotypes where the PRS association was significant after correction, but the effect size was small enough to still be considered significantly equivalent. For example, tobacco use disorder is the most significant result from the TOST test (p = 6.11×10^−64^) reflecting the small effect size of the PRS association (variance explained = 0.054%) but since it’s a common phenotype it is significant after multiple test correction in the PRS association (p = 2.43×10^−7^). This group also included Type 2 diabetes where the schizophrenia PRS was significant but associated with decreased risk. Requiring that the PRS association was not significant after multiple test correction yielded 25 phenotypes, of which 15 had a PRS association p-value > 0.05. Put another way these 15 phenotypes were significantly comorbid with schizophrenia and lacked any discernable difference in schizophrenia PRS between cases and controls despite being well-powered to detect one. This set was enriched for those with prior evidence of non-genetic causes including from medication (e.g., “torsion dystonia”, “convulsions”, “tachycardia”) and lifestyle implications of schizophrenia including reduced hygiene (“diseases of the nail”) or smoking (“bronchitis”).^58–67^

**Table 2:**
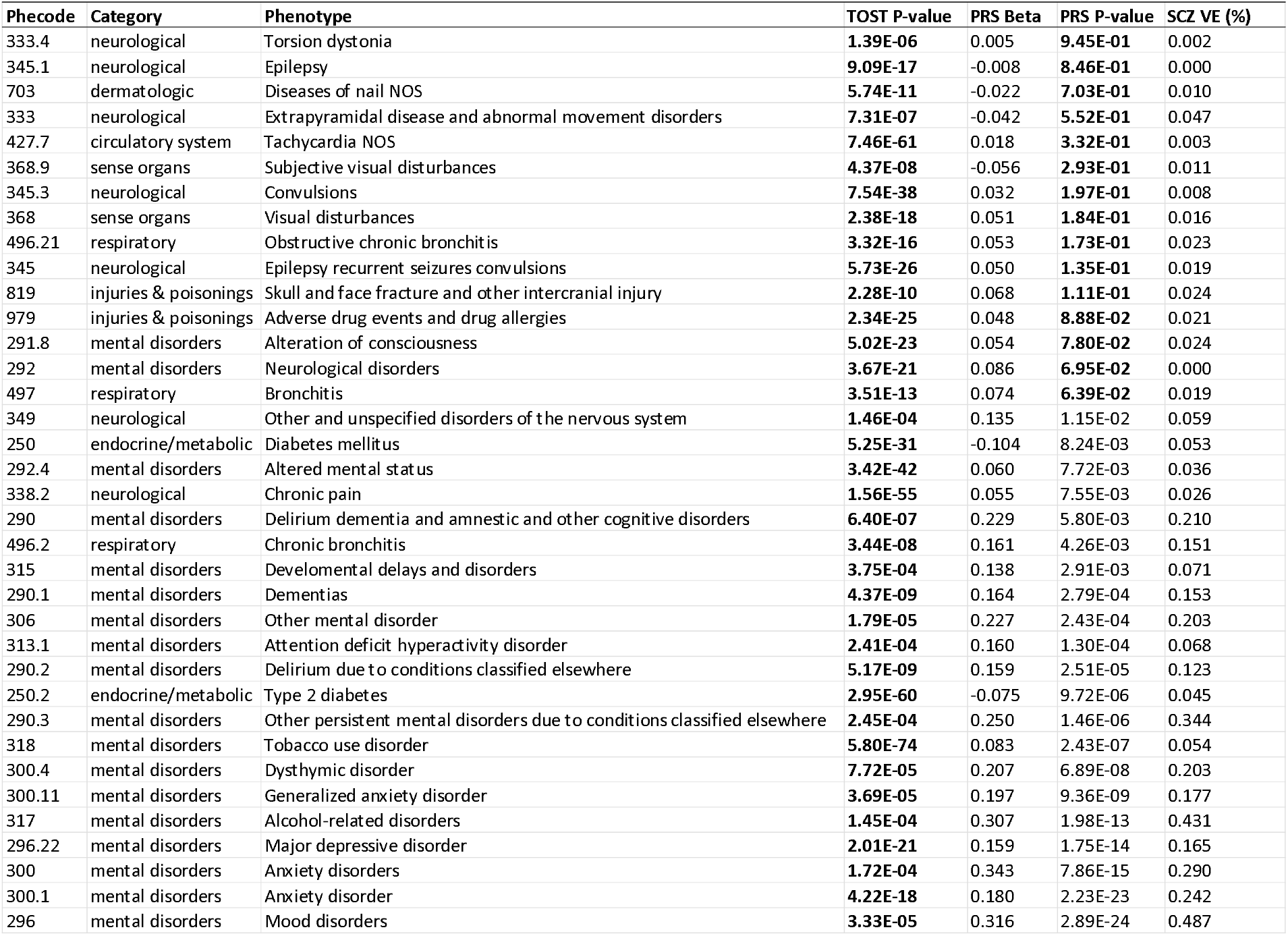
Significant TOST results (p < 0.00065) with non-significant SCZ PRS (p > 0.05) in bold.

| Phecode | Category | Phenotype | TOST P-value | PRS Beta | PRS P-value | SCZ VE (%) |
| --- | --- | --- | --- | --- | --- | --- |
| 333.4 | neurological | Torsion dystonia | **1.39E-06** | 0.005 | **9.45E-01** | 0.002 |
| 345.1 | neurological | Epilepsy | **9.09E-17** | -0.008 | **8.46E-01** | 0.000 |
| 703 | dermatologic | Diseases of nail NOS | **5.74E-11** | -0.022 | **7.03E-01** | 0.010 |
| 333 | neurological | Extrapyramidal disease and abnormal movement disorders | **7.31E-07** | -0.042 | **5.52E-01** | 0.047 |
| 427.7 | circulatory system | Tachycardia NOS | **7.46E-61** | 0.018 | **3.32E-01** | 0.003 |
| 368.9 | sense organs | Subjective visual disturbances | **4.37E-08** | -0.056 | **2.93E-01** | 0.011 |
| 345.3 | neurological | Convulsions | **7.54E-38** | 0.032 | **1.97E-01** | 0.008 |
| 368 | sense organs | Visual disturbances | **2.38E-18** | 0.051 | **1.84E-01** | 0.016 |
| 496.21 | respiratory | Obstructive chronic bronchitis | **3.32E-16** | 0.053 | **1.73E-01** | 0.023 |
| 345 | neurological | Epilepsy recurrent seizures convulsions | **5.73E-26** | 0.050 | **1.35E-01** | 0.019 |
| 819 | injuries & poisonings | Skull and face fracture and other intracranial injury | **2.28E-10** | 0.068 | **1.11E-01** | 0.024 |
| 979 | injuries & poisonings | Adverse drug events and drug allergies | **2.34E-25** | 0.048 | **8.88E-02** | 0.021 |
| 291.8 | mental disorders | Alteration of consciousness | **5.02E-23** | 0.054 | **7.80E-02** | 0.024 |
| 292 | mental disorders | Neurological disorders | **3.67E-21** | 0.086 | **6.95E-02** | 0.000 |
| 497 | respiratory | Bronchitis | **3.51E-13** | 0.074 | **6.39E-02** | 0.019 |
| 349 | neurological | Other and unspecified disorders of the nervous system | **1.46E-04** | 0.135 | 1.15E-02 | 0.059 |
| 250 | endocrine/metabolic | Diabetes mellitus | **5.25E-31** | -0.104 | 8.24E-03 | 0.053 |
| 292.4 | mental disorders | Altered mental status | **3.42E-42** | 0.060 | 7.72E-03 | 0.036 |
| 338.2 | neurological | Chronic pain | **1.56E-55** | 0.055 | 7.55E-03 | 0.026 |
| 290 | mental disorders | Delirium dementia and amnestic and other cognitive disorders | **6.40E-07** | 0.229 | 5.80E-03 | 0.210 |
| 496.2 | respiratory | Chronic bronchitis | **3.44E-08** | 0.161 | 4.26E-03 | 0.151 |
| 315 | mental disorders | Developmental delays and disorders | **3.75E-04** | 0.138 | 2.91E-03 | 0.071 |
| 290.1 | mental disorders | Dementias | **4.37E-09** | 0.164 | 2.79E-04 | 0.153 |
| 306 | mental disorders | Other mental disorder | **1.79E-05** | 0.227 | 2.43E-04 | 0.203 |
| 313.1 | mental disorders | Attention deficit hyperactivity disorder | **2.41E-04** | 0.160 | 1.30E-04 | 0.068 |
| 290.2 | mental disorders | Delirium due to conditions classified elsewhere | **5.17E-09** | 0.159 | 2.51E-05 | 0.123 |
| 250.2 | endocrine/metabolic | Type 2 diabetes | **2.95E-60** | -0.075 | 9.72E-06 | 0.045 |
| 290.3 | mental disorders | Other persistent mental disorders due to conditions classified elsewhere | **2.45E-04** | 0.250 | 1.46E-06 | 0.344 |
| 318 | mental disorders | Tobacco use disorder | **5.80E-74** | 0.083 | 2.43E-07 | 0.054 |
| 300.4 | mental disorders | Dysthymic disorder | **7.72E-05** | 0.207 | 6.89E-08 | 0.203 |
| 300.11 | mental disorders | Generalized anxiety disorder | **3.69E-05** | 0.197 | 9.36E-09 | 0.177 |
| 317 | mental disorders | Alcohol-related disorders | **1.45E-04** | 0.307 | 1.98E-13 | 0.431 |
| 296.22 | mental disorders | Major depressive disorder | **2.01E-21** | 0.159 | 1.75E-14 | 0.165 |
| 300 | mental disorders | Anxiety disorders | **1.72E-04** | 0.343 | 7.86E-15 | 0.290 |
| 300.1 | mental disorders | Anxiety disorder | **4.22E-18** | 0.180 | 2.23E-23 | 0.242 |
| 296 | mental disorders | Mood disorders | **3.33E-05** | 0.316 | 2.89E-24 | 0.487 |

## Discussion

Comorbidities are common among psychiatric patients,^68^ and have been associated with poor outcomes often requiring lengthy hospital admissions and additional monitoring from one or multiple specialists.^69^ Here, we sought to identify schizophrenia comorbidities that are largely unrelated to shared genetic influences leaving other causes that might be more modifiable (e.g. treatment, environment, etc.). We calculated phenome-wide comorbidity using EHR data from two major healthcare systems demonstrating consistency across institutions and identifying 77 significant comorbidities for schizophrenia. These EHR-based comorbidities had significant over-representation in the literature, supporting their validity. When comparing the EHR-based comorbidities to the phenotypes associated with schizophrenia PRS, we identified phenotypes that were significantly comorbid but lacked evidence of shared genetic influences. These results were enriched for phenotypes linked to other causes demonstrating value of comparing comorbidities and genetic associations to identify comorbid phenotypes that might have modifiable causes.

Our EHR-based comorbidity profiles of schizophrenia showed highly significant consistency across two independent healthcare institutions demonstrating the overall robustness of our comorbidity measure. Among schizophrenia comorbidities previously described in multiple studies, we identified 23 of the 55 conditions, a proportion significantly greater than expected by chance. These included 16 of the 17 psychiatric conditions previously reported and 7 of the 38 medical comorbidities. The difference in overlap of the EHR-based comorbidities with the literature between the psychiatric and medical phenotypes is potentially a product of several causes. These include the temporal and specialized care of patients documented in the EHR. Most patients do not receive care for all conditions at the same institution throughout their lives. Since care was provided for schizophrenia, it is more likely that care included psychiatric care which documented the other comorbid psychiatric conditions. However, the extent to which other medical care was provided and for what duration is highly variable and could reduce the power to identify comorbid conditions that occur much later in life or through providers seen at different institutions. Further, our comorbidities are calculated within a hospital population with high comorbidity overall, potentially making it harder to see significant comorbidity for highly prevalent medical conditions such as heart disease and obesity. Nevertheless, psychiatric and medical comorbidities show significant enrichment independently. We observed 47 EHR-based comorbidities that lacked support in the literature presenting an interesting set of comorbidities to follow up further.

The frequent co-occurrence of phenotypes is often matched by shared underlying genetic risk, but there are exceptions. Factors such as unhealthy behavior, environment, and adverse treatment effects can influence comorbidities.^38^ Under these conditions, comorbidities might present without any genetic overlap indicating an opportunity to identify a modifiable cause. Here, we leveraged EHR-based comorbidities with PRS association to identify those exact scenarios for schizophrenia. That is, where schizophrenia significantly co-occurred with another phenotype but there was a significant lack of difference in PRS distributions between cases and controls. We identified 15 phenotypes without even a nominally significant PRS association including “adverse drug events”, as well as “recurrent seizures”, “extrapyramidal disease and abnormal movement disorder”, “tachycardia”, and “torsion dystonia” which have all been previously linked to antipsychotic treatment^58–60,62–64^, fitting our hypothesis. Environmental influences are also apparent such as for “bronchitis” which is influenced by the high rate of smoking in patients with schizophrenia.^65,66^ These results provide validation that this approach is finding comorbidities with other, potentially modifiable, non-common genetic causes. Importantly, there is a set of phenotypes with highly significant equivalence and significant PRS association. This is a product of well-powered PRS analyses but very small effect sizes. Even for the most significant PRS associations among this set, the effect sizes are quite small (variance explained < 0.5%) potentially pointing to non-genetic causes as primary drivers of comorbidities. This set includes several interesting phenotypes such as “tobacco use disorder,” “diabetes” and “dementias” where prior work has implicated genetic sharing and similar etiologies.^70–75^ Our results would suggest that the genetic risk of schizophrenia is not the primary contributing factor to these comorbidities supporting other potential hypotheses and with potential for intervention. In fact, these results indicate that genetic risk of schizophrenia decreases risk of diabetes despite very high comorbidity providing an even stronger case for other contributors such as medication.^35,76^

There are several limitations of this work that should be noted. The first is the use of phecodes as the diagnostic measure for schizophrenia and its comorbidities. ICD codes are designed for clinical documentation and billing purposes, not for research. To reduce the potential of misclassification, we used phecodes that group related ICD codes together, and we required cases to have at least two phecodes recorded on independent dates. However, these codes can vary by clinical practice and since they are captured over time also represent the evolution of a patient’s clinical presentation. For example, bipolar disorder and schizophrenia are clinically independent diagnoses based on the DSM-5, but bipolar disorder is the most significant schizophrenia comorbidity across both sites representing misdiagnosis during clinical presentation over time. Importantly, the strength of comorbidity using these codes was highly consistent across the two institutions. While our approach identifies phenotypes fitting with our expectations, it does not explicitly point to whether those comorbid phenotypes occur before or after diagnosis of schizophrenia and therefore provide insight into what might be potential causes. Efforts to establish more robust inference about causal pathway are necessary. EHR data can be helpful for this process, but substantial consideration will be needed to account for data fragmentation and incomplete time course.

This work provides substantial evidence of the consistency and robustness of comorbidity defined using EHR data based on comparison across multiple healthcare institutions, validation from the literature and consistency with genetic association. We present an approach to intersect EHR-based comorbidities with PRS-based genetic association to identify comorbidities without genetic overlap. These phenotypes present potentially modifiable outcomes driven by other factors such as behavior, treatment, or environment. In schizophrenia, we show multiple examples of this approach working and highlight other phenotypes worthy of further work in identifying the causal path. Overall, this work provides a path to better understanding comorbidities which might be more easily modified to improve outcomes.

## Data Availability

All data produced in the present work are contained in the manuscript. Sharing of individual level data is restricted.

## Acknowledgments

The dataset(s) used for the analyses described were obtained from Vanderbilt University Medical Center’s BioVU, which is supported by institutional funding, private agencies, and federal grants. These include NIH funded Shared Instrumentation Grant S10OD017985, S10RR025141, and S10OD025092; CTSA grants UL1TR002243, UL1TR000445, and UL1RR024975. Genomic data are also supported by investigator-led projects that include U01HG004798, R01NS032830, RC2GM092618, P50GM115305, U01HG006378, U19HL065962, R01HD074711; and additional funding sources listed at https://victr.vanderbilt.edu/pub/biovu/. NS and YX were supported by the Vanderbilt University Department of Biostatistics Development Award. TV was supported by the Human Genetics Training Grant 5T32GM080178-14. KWC was supported in part by funding from the National Institute of Mental Health (K08MH127413) and a NARSAD Brain and Behavior Foundation Young Investigator Award.

## Notes

### Competing Interest Statement

The authors have declared no competing interest.

### Funding Statement

Human Genetics Training Grant 5T32GM080178-14.
NIH funded Shared Instrumentation Grant S10OD017985, S10RR025141, and S10OD025092
CTSA grants UL1TR002243, UL1TR000445, and UL1RR024975. Genomic data are also supported by investigator-led projects that include U01HG004798, R01NS032830, RC2GM092618, P50GM115305, U01HG006378, U19HL065962, R01HD074711. NS and YX are supported by the Vanderbilt University Department of Biostatistics Development Award. KWC was supported in part by funding from the National Institute of Mental Health (K08MH127413) and a NARSAD Brain and Behavior Foundation Young Investigator Award.

### Author Declarations

IRB of Vanderbilt University gave ethical approval for this work under IRB# 172041.

