## supplementary_figures for "Identifying modifiable comorbidities of schizophrenia by integrating electronic health records and polygenic risk"

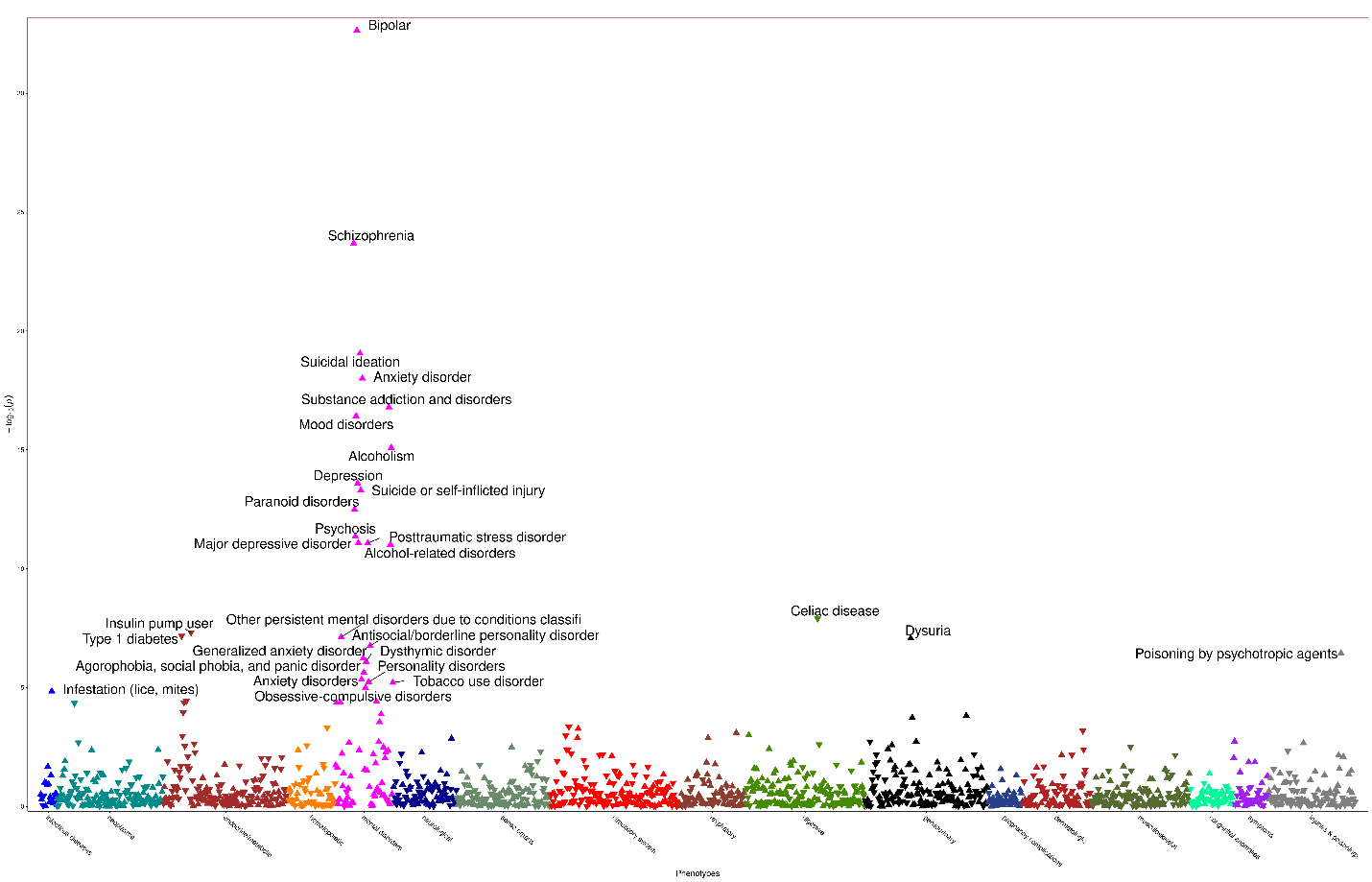

**Supplementary Figure 1: Bonferroni significant EHR associations at VUMC only.** The x-axis is phecodes grouped by category. The y-axis is the negative log10 of the p-values for the phenotypes associated with schizophrenia PRS. No phenotypes were significantly associated. Triangles represent phenotypes with either positive (upright triangle) or negative (inverted triangle) direction of effect.

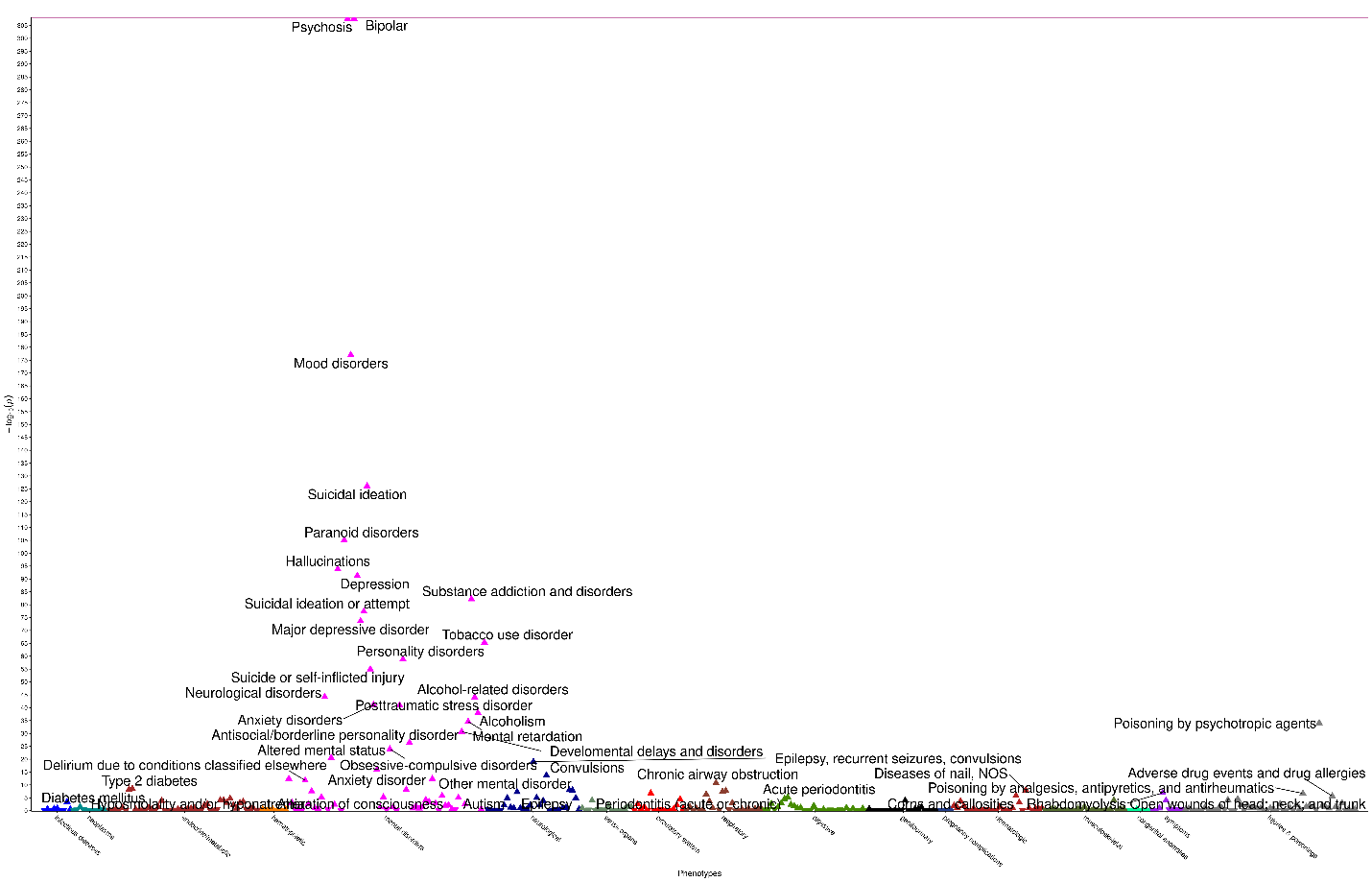

**Supplementary Figure 2: Bonferroni significant EHR associations at MGB only.** The x-axis is phecodes grouped by category. The y-axis is the negative log10 of the p-values for the phenotypes associated with schizophrenia PRS. No phenotypes were significantly associated. Triangles represent phenotypes with either positive (upright triangle) or negative (inverted triangle) direction of effect.

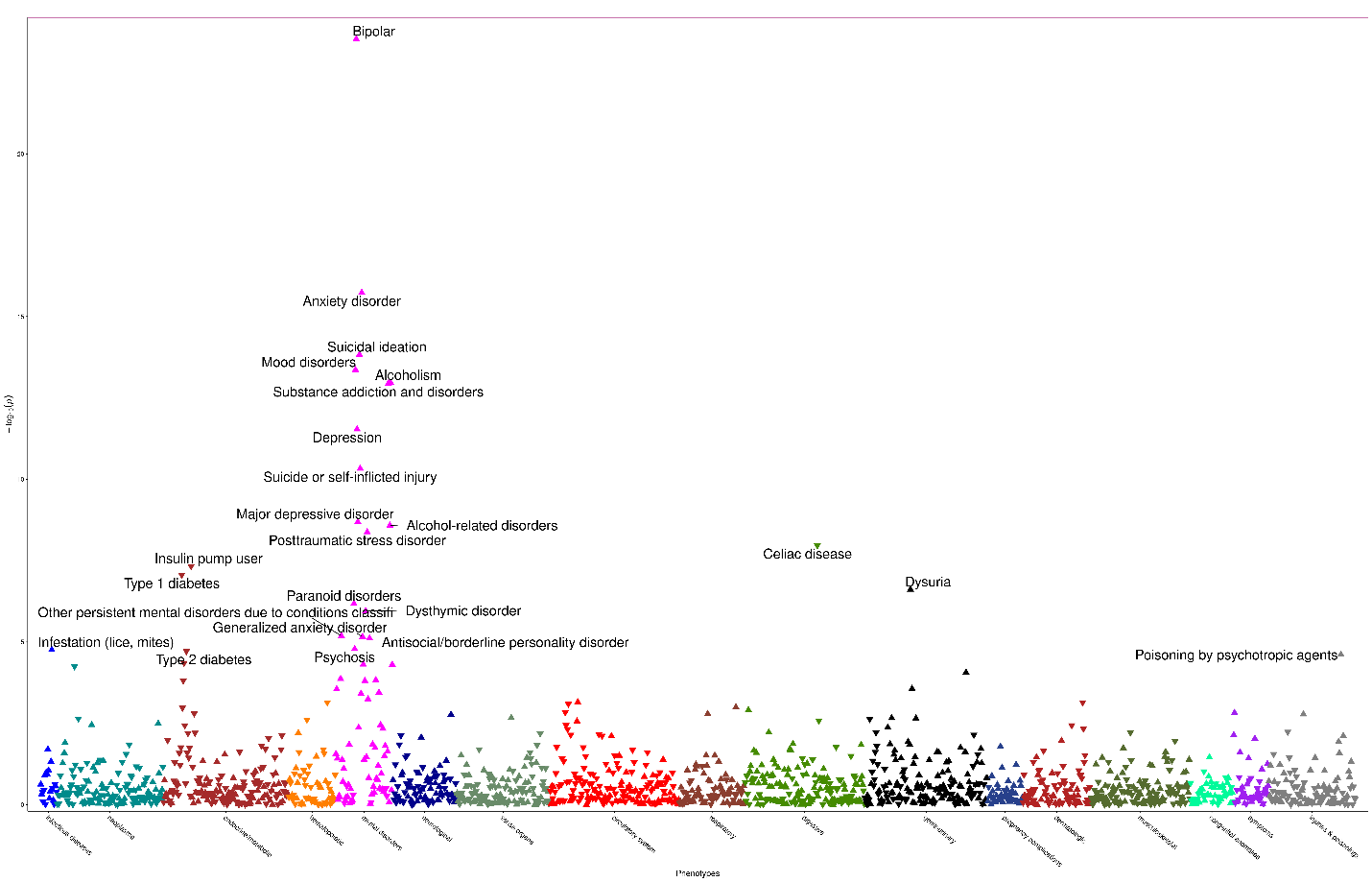

**Supplementary Figure 3: Bonferroni significant SCZ PRS associations at VUMC only.** The x-axis is phecodes grouped by category. The y-axis is the negative log10 of the p-values for the phenotypes associated with schizophrenia PRS. No phenotypes were significantly associated. Triangles represent phenotypes with either positive (upright triangle) or negative (inverted triangle) direction of effect.

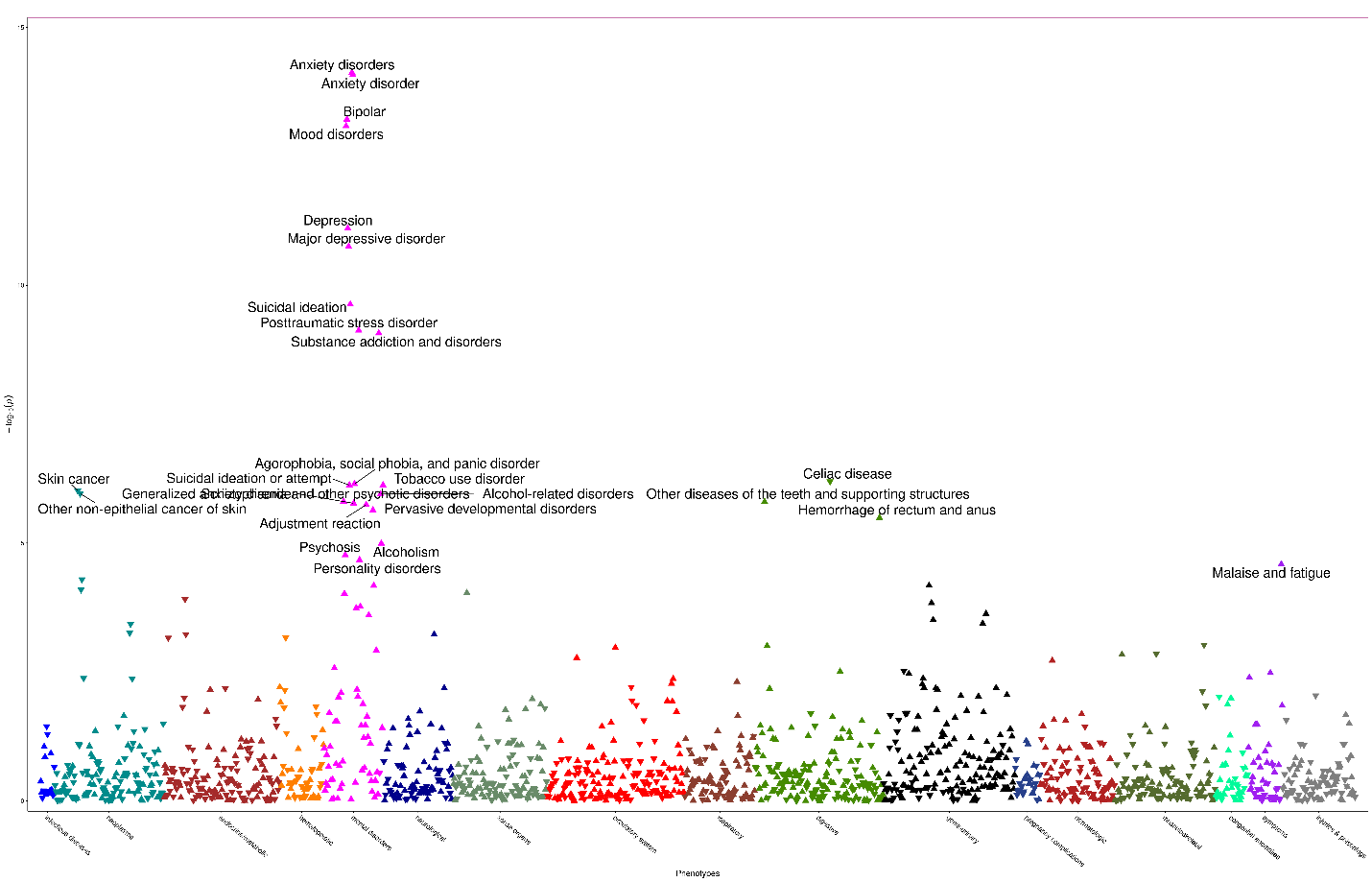

**Supplementary Figure 4: Bonferroni significant SCZ PRS associations at MGB only.** The x-axis is phecodes grouped by category. The y-axis is the negative log10 of the p-values for the phenotypes associated with schizophrenia PRS. No phenotypes were significantly associated. Triangles represent phenotypes with either positive (upright triangle) or negative (inverted triangle) direction of effect.

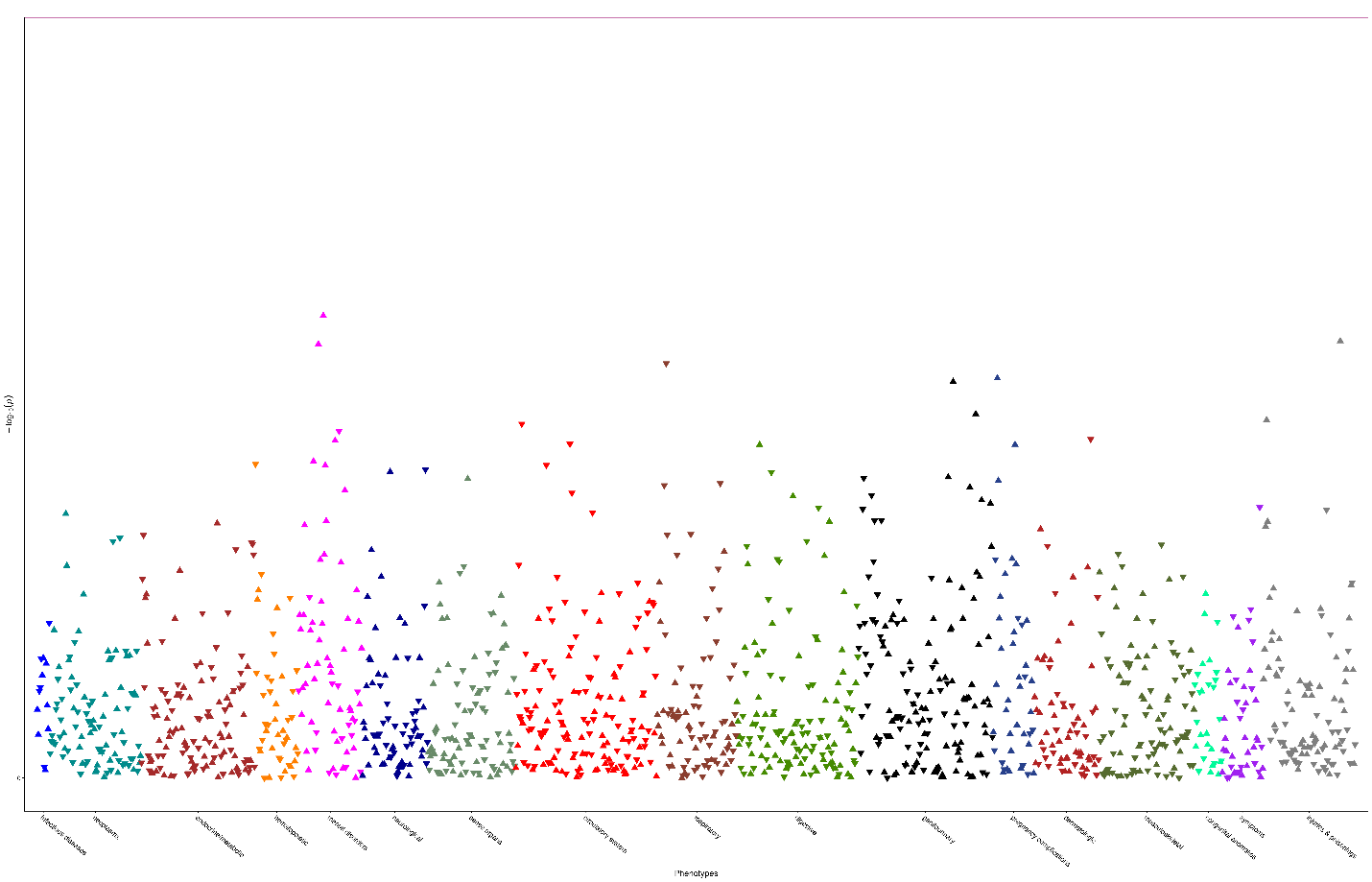

**Supplementary Figure 5: Schizophrenia PRS associations at VUMC for AFR ancestry.** The x-axis is phecodes grouped by category. The y-axis is the negative log10 of the p-values for the phenotypes associated with schizophrenia PRS. No phenotypes were significantly associated. Triangles represent phenotypes with either positive (upright triangle) or negative (inverted triangle) direction of effect.

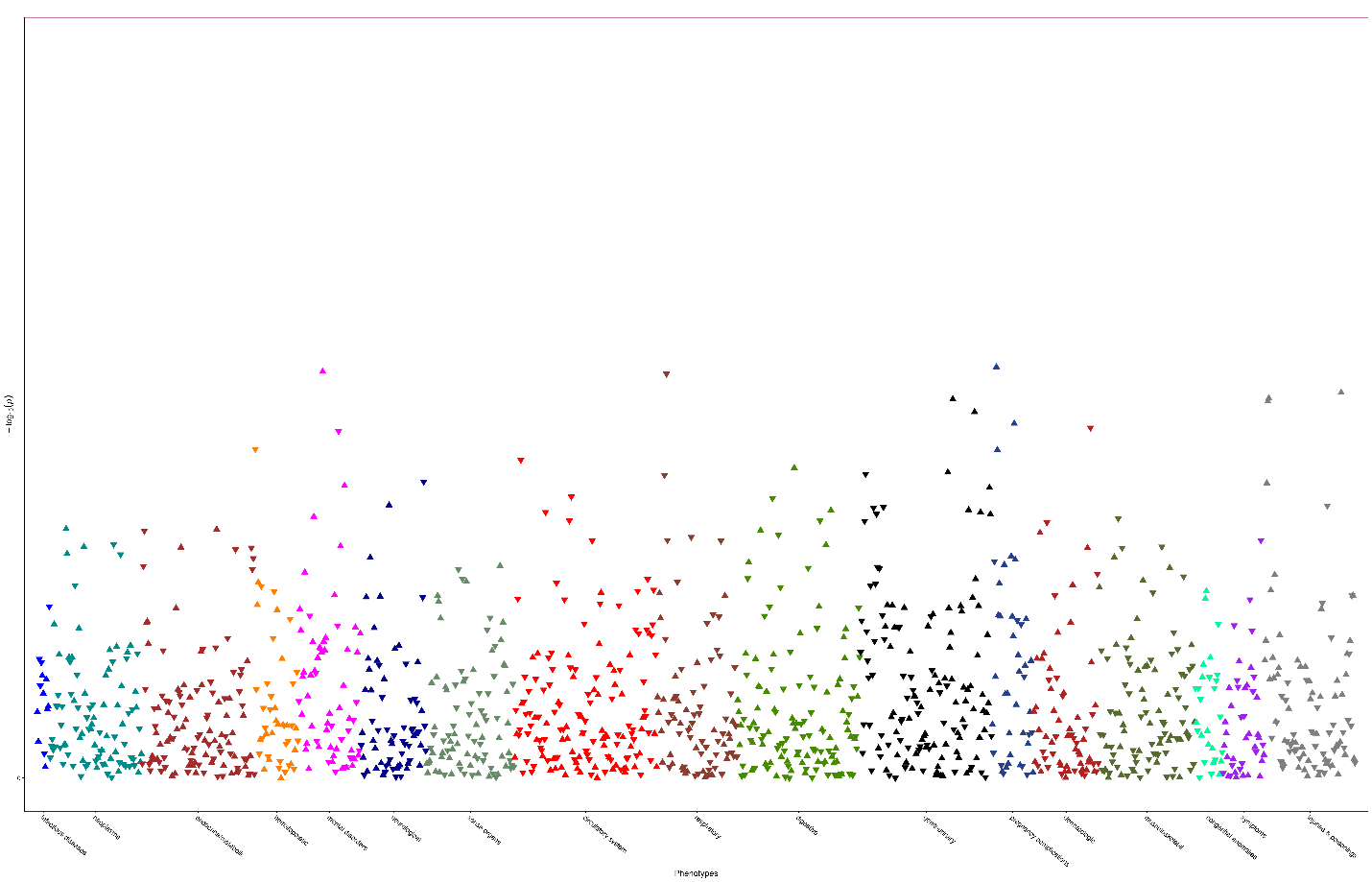

**Supplementary Figure 6: Schizophrenia PRS associations at VUMC for AFR ancestry with schizophrenia patients excluded.** The x-axis is phecodes grouped by category. The y-axis is the negative log10 of the p-values for the phenotypes associated with schizophrenia PRS. No phenotypes were significantly associated. Triangles represent phenotypes with either positive (upright triangle) or negative (inverted triangle) direction of effect.
