## supplementary_methods for "Identifying modifiable comorbidities of schizophrenia by integrating electronic health records and polygenic risk"

**Genotype sample description, quality control (QC) at VUMC**

Samples were genotyped on the Infinium expanded multi-ethnic genotyping array (MEGAEx)^1^ from Vanderbilt’s biobank, BioVU, which initially contained 2,038,233 single nucleotide polymorphisms (SNPs) and 94,474 people. At first, we excluded the SNPs with missing rate >= 0.05 and the positional duplicate SNPs with incompatible alleles. Then we excluded individuals with call rate < 98%, sex discordance, excess heterozygosity rate within each self-reported ancestry, or potentially cross-contaminated individuals [proportion identity-by-descent (IBD) > 0.8 between different individual IDs]. We next excluded SNPs with missing rate > 0.02, minor allele frequency (MAF) < 0.01, Hardy-Weinberg equilibrium (HWE) test P value ≤ 1x10^–6^ within self-reported ancestry of AFR or EUR, or MAF < 0.01 within the whole sample. Thus, there were 90,313 individuals which were kept with a set of high-quality autosomal SNPs (N=887,250).

We downloaded 1000 Genomes phase 3 (1000 GP3) data^2^, which consists of 2,504 unrelated samples from 5 super populations African (AFR), Admixed American (AMR), East Asian (EAS), European (EUR), South Asian (SAS). Next, we extracted 887,250 QC-ed SNPs (no INDELs) that are genotyped on the BioVU MEGAEx array and merged it with 1000 GP3 data after removing C/G and A/T SNPs to avoid unresolvable strand mismatches in MEGA samples. Regions with known high linkage disequilibrium (LD) were excluded and the common variants were then pruned (r^2^ < 0.05) using PLINK 1.9 (–indep-pairwise 1000 50 0.05) to yield 71,339 SNPs in relative linkage equilibrium for ancestry analyses.^3^ The flashpca version 2.0 was used to perform PCA to generate 2 top genetic PCs.^4^ By using K nearest neighbors (KNN) clustering with k = 5, we inferred MEGA samples' ancestries based on the top 2 PCs from the 1000 GP3 and MEGA samples. We treated 1000 GP3 samples as the training set and MEGA samples as the testing set. For each MEGA individual, we calculated its Euclidean distance from each individual in the training set. Next, we sorted the distance, and found the 5 nearest neighbors from the 1000 GP3 based on the 5-th minimum distances for each MEGA individual. Based on whether all of the nearest neighbors were from the same population, we inferred the detected MEGA individual's ancestry. (If they are from more than one super population, we assigned the individual's ancestry as admixed one.) Among the 90,313 MEGA individuals, 87,558 (96.95%) were assigned to five homogeneous super-populations, i.e. AFR=13,808, AMR=2,446, EUR=70,473, EAS=441, SAS=390.^5^ The final set used in our main analyses was the EUR only subgroup.

Prior to imputation, the SNP position, alleles, Ref/Alt assignments and frequency differences were checked by comparison with the Haplotype Reference Consortium (HRC) panel (Version r1.1 2016)^6^ in build GRCh37^7^ using McCarthy Group Tools (https://www.well.ox.ac.uk/~wrayner/tools/). SNPs with inconsistent alleles, with > 0.2 allele frequency difference, or not in the reference panel were removed. Phasing and imputation were then performed using a standard pipeline on the Michigan Imputation Server (MIS).^8^ The total QC-ed SNP data was divided into five batches. Phasing was performed using Eagle version v2.4.^6^ The HRC reference panel in build GRCh37 was selected using mixed population.^6,7^ The genotype probabilities in post-imputed data were converted to hard-call genotypes using PLINK2 (hard-call >= 0.1).^9^ SNPs were removed with imputation info score in any of the batches < 0.3, position duplicates, missing genotype rate > 0.02, or multi-allelic states (>2). For autosomal data, we reused a series of QC filtering steps, excluding SNPs with MAF < 0.005, HWE test P value < 1x10^–6^, and removing the individuals with missing rate ≥ 0.02, excess heterozygosity rate over 3* interquartile range (IQR) of the upper heterozygosity quartile (Q3) for each subset.

**Genotype sample description, quality control (QC) at MGB**

Mass General Brigham Biobank (MGBB) samples were genotyped using the Illumina Multi-Ethnic Global array with hg19 coordinates.^1,7^ MGBB initially contained 1.7 million SNPs and 36,424 individuals. Variant-level quality control filters removed variants with a call rate < 98%, as well as those that were duplicated across batches, monomorphic, not confidently mapped to a genomic location, or showed an association with genotyping batch. Sample-level quality control filters removed individuals with a call rate < 98%, MAF < 0.05, strand ambiguous SNPs and long-range LD regions (chr6:25-35Mb; chr8:7-13Mb inversion).

Principal components of ancestry were calculated in the 1000 GP3 reference panel^2^ and subsequently projected onto the MGBB dataset, where a Random Forest classifier was used to assign ancestral group membership for those with a prediction probability > 90%. Using the assigned ancestral group membership, we identified individuals with EUR ancestry. Within the European samples, sample-level quality control filters removed samples with excessive autosomal heterozygosity (±3 standard deviations from the mean), related individuals (pi-hat < 0.2), or discrepant self-reported and genetically inferred sex. Variant-level quality control filters removed SNPs with a call rate < 98%, HWE test P value < 1x10^–10^ and retained only autosomal SNPs (exclude indels and monomorphic SNPs).

The MIS^8^ was then used to impute missing genotypes with the HRC^6^ dataset serving as the reference panel. Imputed genotype dosages were converted to hard-call format and subjected to further quality control, where SNPs were removed if they exhibited poor imputation quality (INFO < .80), call rate < 98%, MAF < 0.01, or HWE test P value < 1x10^–10^. These procedures yielded a final analytic sample of 25,698 individuals in the MGBB with a set of high-quality autosomal SNPs (N=909,229).

**AFR ancestry schizophrenia PRS calculation and PheWAS (VUMC only)**

PRS were calculated using the Python based program PRS-CS and the 1000 Genomes Project African cohort to represent the LD structure between SNPs.^2,10^ The SNP effect sizes from the Psychiatric Genomic Consortium’s (PGC) latest available genome-wide association study (GWAS) for schizophrenia were used to calculate the PRS.^11^ These results came from a meta-analysis of 9 cohorts that included 6,152 cases and 3,918 controls of African ancestries.^11^ The scaling parameter, phi, was set to 1e-2 to represent the polygenic architecture of schizophrenia. The PRS-CS generated posterior effects for SNPs were then used to calculate PRS for each cohort using the PLINK–score flag with sum as a modification to output SCORESUM instead of SCORE.^3^ As with the EUR sample, PRS was calculated with schizophrenia patients excluded from the PheWAS analyses to exclude association as a byproduct of comorbidity.

Since we estimated PRS for all those genotyped without a schizophrenia diagnosis, there were 1,406 phecodes out of the possible 1,866 with at least one person as a case for the AFR cohort. For each of these 1,406 phecodes, we performed a logistic regression testing its association with the schizophrenia PRS including covariates of sex, current age, record length in days, and the first 10 principal components (PCs) for African ancestry. As with EUR ancestry, at least two instances of the phecode had to be present to be considered a case and phecodes listed as exclusions were removed from the controls.

**References**

1.	Multi-Ethnic Genotyping Array Consortium. https://www.illumina.com/science/consortia/human-consortia/multi-ethnic-genotyping-consortium.html.

2.	1000 Genomes | A Deep Catalog of Human Genetic Variation. https://www.internationalgenome.org/home.

3.	Allelic scoring - PLINK 1.9. https://www.cog-genomics.org/plink/1.9/score.

4.	Abraham, G., Qiu, Y. & Inouye, M. FlashPCA2: principal component analysis of Biobank-scale genotype datasets. *Bioinformatics* **33**, 2776–2778 (2017).

5.	NHLBI Trans-Omics for Precision Medicine (TOPMed) Consortium *et al.* Sequencing of 53,831 diverse genomes from the NHLBI TOPMed Program. *Nature* **590**, 290–299 (2021).

6.	Loh, P.-R. *et al.* Reference-based phasing using the Haplotype Reference Consortium panel. *Nat Genet* **48**, 1443–1448 (2016).

7.	GRCh37 - hg19 - Genome - Assembly - NCBI. https://www.ncbi.nlm.nih.gov/assembly/GCF_000001405.13/.

8.	Das, S. *et al.* Next-generation genotype imputation service and methods. *Nat Genet* **48**, 1284–1287 (2016).

9.	PLINK. https://zzz.bwh.harvard.edu/plink/plink2.shtml.

10.	Ge, T., Chen, C.-Y., Ni, Y., Feng, Y.-C. A. & Smoller, J. W. Polygenic prediction via Bayesian regression and continuous shrinkage priors. *Nat Commun* **10**, 1776 (2019).

11.	Trubetskoy, V. *et al.* Mapping genomic loci implicates genes and synaptic biology in schizophrenia. *Nature* **604**, 502–508 (2022).
